# Pathogenic MYRF ICA Domain Variant is Aberrantly Trafficked in the RPE and Results in Isolated Nanophthalmos

**DOI:** 10.64898/2026.11.08.26367379

**Authors:** Gabrielle M. Rozumek, Rachel S. Lim, William Presley, Bridget Blevins, Bin Guan, John Y.S. Han, Ehsan Ullah, Amelia Naik, Tara E. Hahn, Rodrigo Young, Sayoko E. Moroi, Robert B. Hufnagel, Jason M.L. Miller, Lev Prasov

**Affiliations:** Department of Ophthalmology and Visual Sciences, University of Michigan, Ann Arbor, MI, USA; Department of Pathology, University of Michigan, Ann Arbor, MI, USA; Department of Human Genetics, University of Michigan, Ann Arbor, MI, USA; Department of Human Genetics, University of Chicago, Chicago, IL, USA; National Eye Institute, National Institutes of Health, Bethesda, MD, USA; Houston Methodist Eye Associates, Houston Methodist, Houston, TX, USA; Institute of Ophthalmology, University College of London, London, UK; Department of Ophthalmology and Visual Sciences, The Ohio State University, Columbus, OH, USA; Center for Integrated Healthcare Research, Kaiser Permanente Hawaii, Honolulu, Hawaii, United States; Cellular and Molecular Biology Program, University of Michigan, Ann Arbor, Michigan, USA

**Author notes:** Author for correspondence: Lev Prasov, MD, PhD, KEC 229, 1000 Wall St., Ann Arbor, MI 48105.

**Keywords:** ICA domain, Myelin regulatory factor (MYRF), Eye growth, Nanophthalmos, retinal pigment epithelium (RPE), Cleavage deficiency, missense, Cardiac-urogenital syndrome, Protein trafficking

## Abstract

**Purpose:** Myelin regulatory factor is a transcription factor essential for RPE development and serves as a critical “GO” signal for eye growth. Pathogenic variants in MYRF manifest in a spectrum of clinical phenotypes ranging from isolated nanophthalmos to severe syndromic disease, in a gene dosage dependent manner. In this study, we aimed to determine the pathogenicity and molecular mechanism of a novel variant in the MYRF ICA domain, c.2083C>T (p.R695C), that leads to isolated nanophthalmos and mild retinal pigment epithelial disruption.

**Methods:** Whole genome sequencing was performed and identified a novel MYRF variant. Pathogenecity and *in silico* modeling were evaluated using the Ensembl VEP tool and PyMOL. Human primary RPE and/or ARPE-19 cells were transduced with FLAG-tagged variant and wildtype MYRF or double-tagged FLAG-MYRF-MYC proteins to evaluate transcriptional activity, localization, and protein trafficking.

**Results:** *In silico* modeling supports variant pathogenicity and altered homotrimer conformation. Functional studies in human primary RPE (hRPE) and/or ARPE-19 cells confirmed that MYRF p.R695C is cleavage deficient, mislocalized to the apical surface of polarized RPE, and demonstrated reduced synergistic transcriptional activity with co-factor SOX10 in HEK293T cells. Protein trafficking studies demonstrated that wildtype MYRF is trafficked to the apical cell surface and plasma membrane and that endogenous MYRF requires ER to Golgi trafficking pathways for proper nuclear localization and cleavage dynamics.

**Conclusions:** Together, these results confirm that p.R695C is pathogenic for nanophthalmos, implicate trafficking as a regulatory mechanism of MYRF in the hRPE, and support that RPE disruption as a feature of MYRF-associated nanophthalmos.

## INTRODUCTION

Proper focusing of light on the retina requires appropriate eye growth during development through two critical periods. The first occurs during embryogenesis ^1,2^ and the second, vision-guided, growth phase occurs postnatally ^3^. Failure of ocular axial elongation results in hyperopia/farsightedness. Nanophthalmos is characterized by an extreme reduction in axial length in the context of an otherwise structurally intact eye, and it can be associated with blinding complications such as glaucoma, retinal degeneration, and retinal detachment ^4^.

Previously, our lab has shown that the transcription factor Myelin Regulatory Factor (MYRF) is a highly and specifically transcription factor and acts as a necessary “GO” signal for eye growth and elongation ^5–7^. It is a type-II transmembrane protein that localizes to the ER lumen that is cleaved to release an N-terminal fragment that translocates into the nucleus and activates gene expression as a functional trimer^8,9^. MYRF contains multiple functional domains, including the highly conserved DNA binding domain (DBD) ^10^, intramolecular chaperone domain (ICA) responsible for cleavage ^11,12^, transmembrane domain (TM) ^9^, and a C-terminal domain (CTD). The DBD, ICA, and TM are highly constrained, and variants therein often result in *MYRF*-related cardiac urogenital syndrome (*MYRF*-CUGS) ^14^. Features include congenital diaphragmatic hernias, heart defects, lung hypoplasia, urogenital abnormalities, and nanophthalmos ^14^. Interestingly, variants within the CTD are associated with a predominantly ocular disease presentation with few or no syndromic features ^6,7,13,15^. Previous studies from our lab and others suggests that 15-19% loss MYRF gene dosage results in isolated ocular disease^7^, whereas 50% loss results in syndromic disease ^10,11,13^.

Previously described substitutions in the DBD and ICA act via haploinsufficiency or dominant-negative mechanisms – including impaired homotrimerization, ^16^, auto-proteolytic cleavage, and/or transcriptional activity – depending on the biochemical effects of specific amino acid substitutions ^10–12^. The *de novo* ICA missense variants *MYRF* p.V679A and R695H result in loss of function due to deficient cleavage ^11^, with the p.R695H identified in a patient with congenital diaphragmatic hernia, a heart defect, and genital anomalies^11^.

Here, we employ *in silico* modeling and *in vitro* functional assays to characterize the pathogenic mechanisms of *MYRF* p.R695C, which was found in a case of isolated nanophthalmos. We also investigate RPE trafficking of endogenous, WT-MYRF – as well as cleavage deficient variants (p.V679A and p.R695C) – in RPE to explore whether tissue-specific processing/functions of MYRF could account for the different phenotypes associated variants impacting the same regions of the protein. Our results support that *MYRF* p.R695C results in nanophthalmos through loss of cleavage competency resulting in reduced transcriptional activation. Furthermore, we provide novel mechanistic insights into how the trafficking of MYRF may impact protein processing and transcriptional activity in RPE.

## RESULTS

### Clinical Features of a Proband with Nanophthalmos

The mixed-ancestry proband presented in her 30s with high hyperopia (+10.0 spherical equivalent refraction [SER] OD, and +13.5 SER OS), short axial length (18.54mm to 18.97mm) (Figure 1A), a shallow anterior chamber featuring plateau iris configuration (Figure 1B), patchy granular subretinal disruption and possible sub-RPE deposits in the macula (Figure 1C), and normal foveal contour (Figure 1D). Follow up near-infrared (NIR) and OCT imaging after 13 years of follow-up showed mild progression of subretinal and sub-RPE irregularities (Figure 1E-G). Clinical course following cataract surgery was notable for uveitis and aqueous misdirection in the left eye, which are both common findings in nanophthalmic eyes ^17^. Systemic history was notable for hypertension, but no cardiac, pulmonary, or urogenital malformations. Available family members (children) were unaffected with normal eye exams.

**Figure 1.**
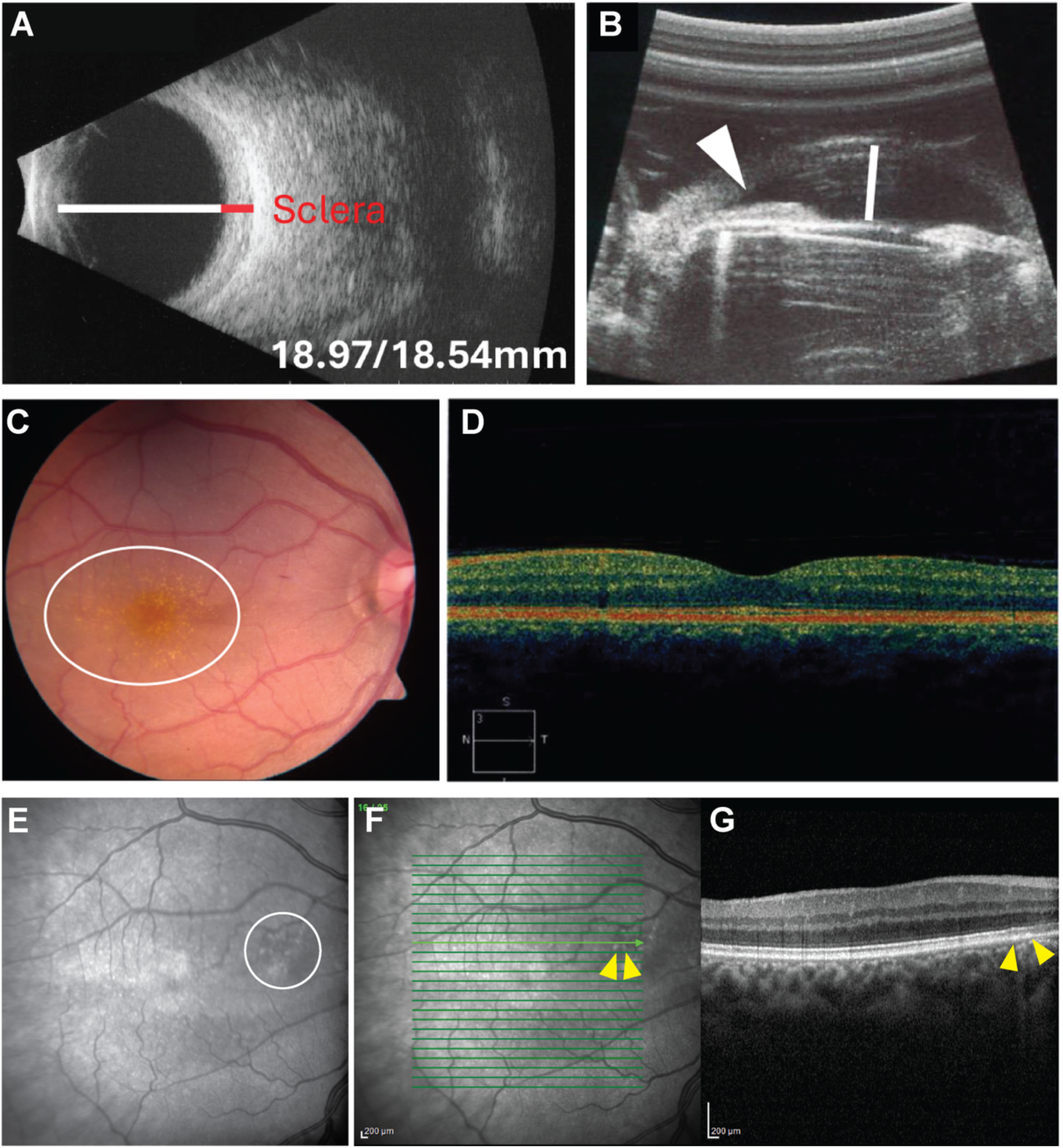
Clinical features of proband carrying R695C-MYRF variant. **(A-D)** Images from initial diagnosis in her 30s show **(A)** Posterior B-scan (OS) showing thickened sclera (red bar) and decreased axial length (white bar). **(B)** Ultrasound biomicroscopy showing shallow anterior chamber (white bar), narrow angle and plateau iris (arrowhead). **(C)** Disc photo (OD) with fine macular drusenoid deposits. **(D)** Optical coherence tomography, OCT, (OS) shows no signs of foveal hypoplasia. **(E-G)** Near-infrared (NIR) and **(G)** SD-OCT images after 13 years follow-up showing subretinal irregularities (yellow arrows, **F-G**). OCT image represents the cross-section from the raster line (green arrow, **F**).

Whole genome sequencing of the proband identified a novel pathogenic missense substitution in *MYRF*, NM_001127392.3 c.2083C>T p.(R695C) and no other variants in known nanophthalmos genes^4^. This variant was absent in two unaffected children, but parents were unavailable to assess *de novo* status. *In silico* predictors suggest that *MYRF* p.R695C is highly conserved (Figure 2A) and deleterious, with scores comparable to those of two known pathogenic and cleavage-deficient ICA variants, p.V679A and p.R695H (Table 1). Furthermore, crystallography-based molecular modeling of the MYRF ICA domain (pdb_00007dc3) ^12^ revealed similar losses of hydrogen bonding stemming from the p.R695C and p.R695H substitution (Figure 2B-D). Together, these findings support that *MYRF* p.R695C results in a conformational change of the ICA domain that could alter cleavage.

**Figure 2.**
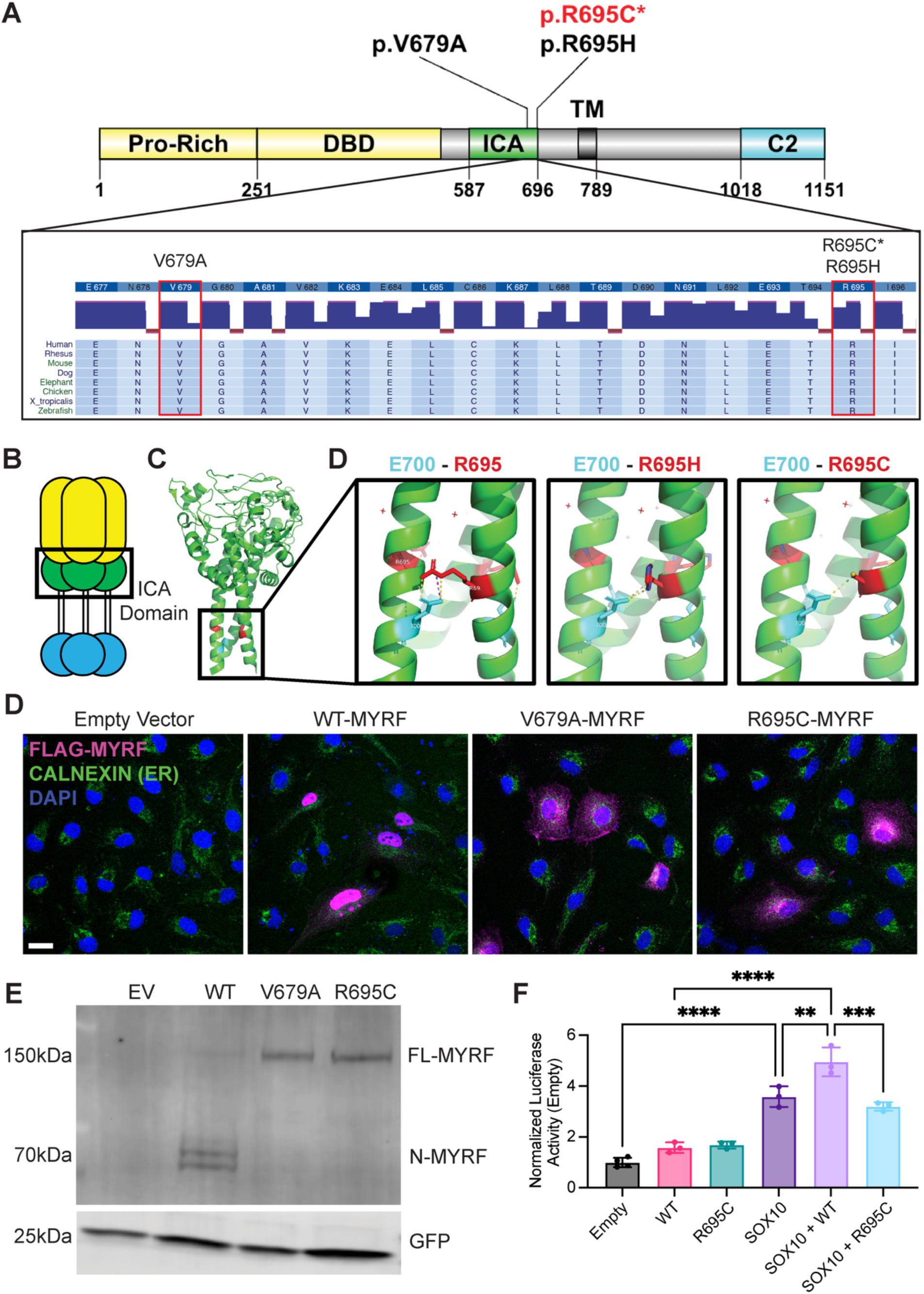
MYRF p.R695C variant disrupts ICA domain conformation and is cleavage deficient leading to reduced transcriptional activity. **(A)** Schematic of MYRF protein and ICA domain location. (B) Crystal structure of ICA domain homotrimer with E700 (highlighted in cyan) and R695 (highlighted in red) residues marked (PBD ID: 7dc3). (C) Structural modeling of wild-type (WT) and known cleavage deficient variant R695H compared to R695C. Note loss of salt bridge and hydrogen bonds in R695H and R695C. (D) ARPE-19 cells transfected with MYRF-R695C variant show absence of nuclear localization and instead cytosolic localization (n=3). Scale bar = 20µm. (E) Western blot of FLAG tagged MYRF constructs shows only full-length (FL) 140kDa band for R695C and V697A, while WT shows N-terminal (N) cleavage product (90kDa), suggesting cleavage deficiency (n=2). (F) Luciferase assays in HEK293T cells using synthetic promoter with MYRF and SOX10 binding sites (dimuc c7c from ^18^ show that the R695C variant lacks synergistic transcriptional activation with SOX10 (n=3, showing one representative graph). Only key statistical comparisons are shown.

**Table 1.** MYRF-R695C Variant is Predicted to be Pathogenic.

| **Substitution** | **Polyphen** | **Allele Frequency** | **CADD** | **REVEL** | **AlphaMissense** | **ACMG Criteria Met** | **ACMG Classification** | **Disease** |
|---|---|---|---|---|---|---|---|---|
| p.V679A | 0.847 | absent | 25.6 | 0.318 | 0.974 | **Supporting:** PP2, PP3<br>**Moderate:** PM2<br>**Strong:** PS3 | Pathogenic | Syndromic |
| p.R695H | 0.952 | 7.15E-07 | 32 | 0.401 | 0.94 | **Supporting:** PP2, PP3<br>**Moderate:** PM2<br>**Strong:** PS3 | Pathogenic | Syndromic |
| p.R695C | 0.998 | absent | 34 | 0.387 | 0.9879 | **Supporting:** PP2, PP3<br>**Moderate:** PM5, PM2<br>**Strong:** PS3 | Pathogenic | Nanophthalmos |

PP = supporting, PM = moderate, PS = strong (American College of Medical Genetics Pathogenic criteria), (Richards et al., 2015)

### R695C Variant Inhibits MYRF Cleavage and Alters Transcriptional Activation

To test for cleavage deficiency, we expressed either WT or variant 3xFLAG-MYRF in ARPE-19 cells. WT-MYRF showed nuclear localization of the N-terminal fragment as expected. Conversely, cleavage-deficient MYRF p.V679A showed a diffuse cytosolic distribution and co-localized with the ER (as labeled with calnexin) (Figure 2D). Interestingly, our MYRF p.R695C variant also showed similar colocalization with nuclear exclusion (Figure 2D). Next, we probed western blots for FLAG-tagged WT, V679A, or R695C MYRF to assess any underlying changes in cleavage dynamics. We observed both full length (140kDa) and N-terminal-cleaved (90kDa) bands for WT-MYRF. However, MYRF p.V679A and p.R695C demonstrated only one robust band at 140kDa corresponding to the full-length protein (Figure 2E). These data further support that MYRF p.R695C is cleavage deficient.

The N-terminal cleavage product of MYRF is important for its ability to activate gene expression. To evaluate the transcriptional activity of MYRF p.R695C, we used established synthetic luciferase plasmids (dimluc c7c) containing SOX10 and MYRF binding sites, as prior studies established that MYRF works in conjunction with SOX10 to activate or repress gene targets in oligodendrocytes ^18^. ARPE-19 cells co-transfected with SOX10 and WT-MYRF expression constructs showed enhanced luciferase activity in comparison to SOX10 alone as previously reported (5.0±0.6 vs. 3.6±0.4 FC) ^18^ (Figure 2F). However, cells co-transfected with SOX10 and R695C-MYRF showed diminished luciferase activity by comparison (3.0±0.2 vs. 5.0±0.0.6 FC) (Figure 2F). Together, our results indicate that the p.R695C substitution leads to disruption of the ICA conformation, cleavage deficiency, and a reduced transcriptional activity, in part through loss of synergistic activation with SOX10.

### Cleavage Deficient Variants Mislocalize Apically in Human Primary RPE (hRPE)

To evaluate whether RPE differentiation state and polarization alter cleavage dynamics of MYRF, we repeated key ARPE-19 experiments in hRPE cultures grown on microporous supports, as described previously ^19^. We transduced mature hRPE cultures (2 months old) with FLAG-tagged WT-, V679A-, or R695C-MYRF lentiviral constructs (Figure 3A). WT-MYRF almost exclusively demonstrated nuclear localization in central optical sections of polarized hRPE, suggesting near complete processing and translocation (Figure 3B). As expected, both MYRF p.V679A and p.R695C were unable to cleave and absent from the nuclei and central optical sections (Figure 3B). However, unlike the results observed in ARPE-19 cells and prior studies using oligodendrocytes ^8,11,16^, cleavage-deficient variants did not co-localize with ER marker calnexin. Instead, we observed that the cleavage-deficient forms of MYRF co-localizing with actin-binding protein phalloidin in the apical microvilli of the RPE (Figure 3B,C). Western blot analysis subsequently supported similar cleavage dynamics of MYRF p.R695C in hRPE as compared to ARPE-19 cells, though a second, unknown band species was observed around 110kDa (Figure 3D). The overall health of hRPE overexpressing WT-, V679A-, or R695C-MYRF was assessed using transepithelial electrical resistance (TEER), a marker of tight junction integrity ^19–21^. TEER was unchanged at 1 week post-transduction suggesting that mislocalization of MYRF to the apical microvilli is not driven by lentiviral induced toxicity. Together, these data suggest that the cell type (oligodendrocyte vs. RPE), differentiation state, and polarization (mature primary RPE vs. ARPE-19) are important factors in the processing and localization of MYRF.

**Figure 3.**
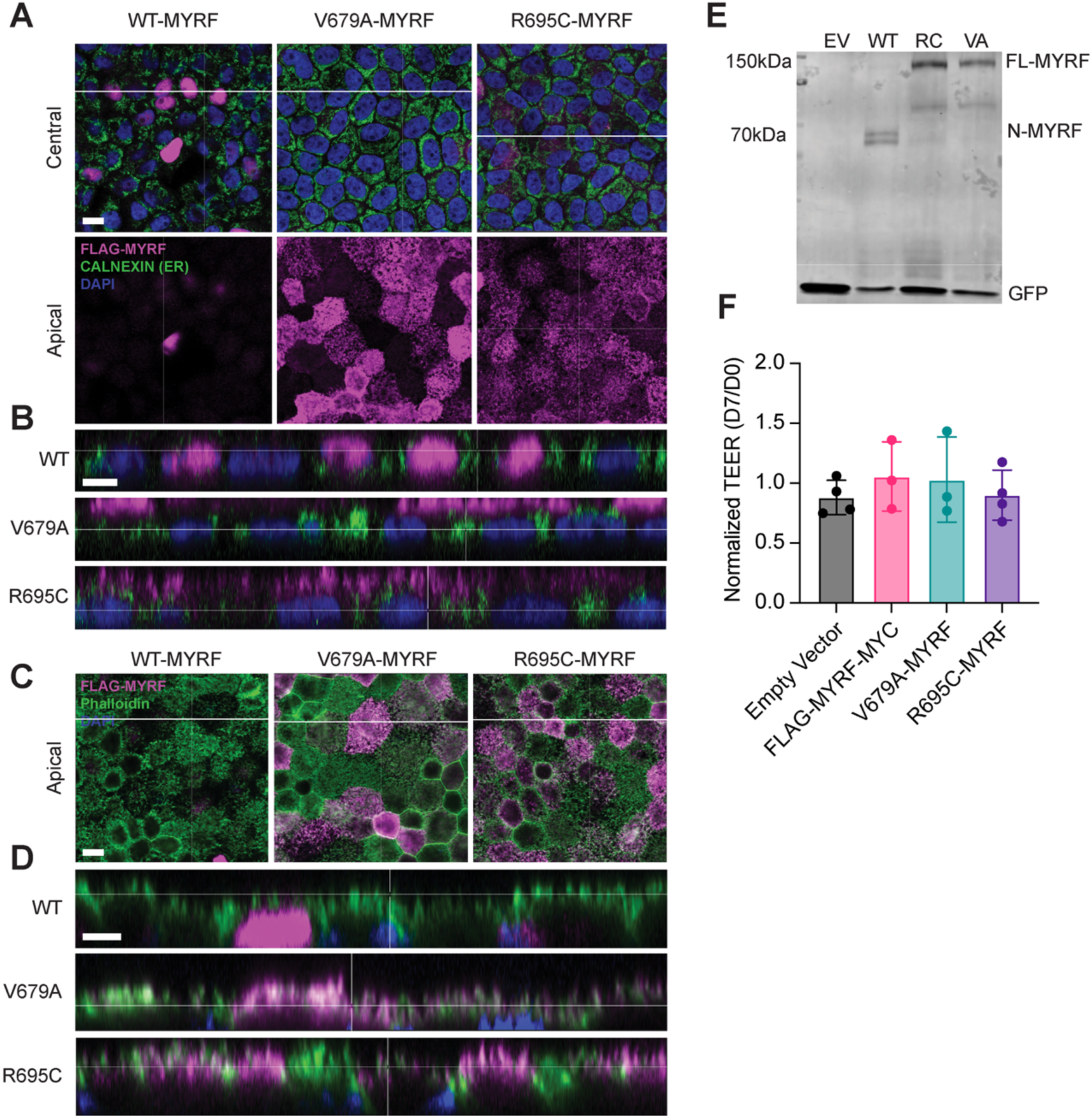
Cleavage deficient variants mislocalize apically in mature hRPE. (A) Central and apical sections from confocal z-stacks and cross sections of hRPE transduced with MYRF variants. FLAG-tagged cleavage deficient variants V679A and R695C are mislocalized to the apical part of the cell in bleached hRPE (A-B). White lines indicate where Z-stack cross section is located. Magnified Z-stack cross sections showing apical mislocalization (B). FLAG-tagged MYRF proteins colocalize with the apical microvilli marker (phalloidin) in unbleached hRPE (C-D). Magnified Z-stack cross sections showing apical mislocalization and colocalization with phalloidin (D). (E) Western blot of hRPE transduced with FLAG tagged MYRF proteins shows R695C is cleavage deficient in mature polarized RPE (n=2-3 donors). (F) Normalized transepithelial electrical resistance (TEER) were not significantly different between conditions (n=3-4 donors). Scale bars = 10µm.

### MYRF is Localized to the Plasma Membrane in hRPE

Given the unexpected apical mislocalization of cleavage-deficient MYRF variants p.V679A and p.R695C, we further investigated the processing of WT MYRF in hRPE. In oligodendrocytes, MYRF is thought to homotrimerize at the ER membrane before undergoing cleavage ^8,11,16^. However, in *C. elegans*, MYRF homolog MYRF-1 is trafficked to the plasma membrane to be cleaved ^22^. To assess the trafficking pattern of WT-MYRF in RPE, we evaluated cellular localization of endogenous MYRF using a MYRF antibody targeting the N-terminus ^23^ and by detecting an overexpressed dual-tagged FLAG-MYRF-MYC protein.

Notably, we observed both nuclear and apical localization of endogenous MYRF (Figure 4 A-C). The nuclear localization can be attributed to the N-terminal cleavage product ^8–10,16,24^ as uncleaved MYRF is excluded from the nucleus (Figure 3) due to transmembrane tethering. In contrast, the apically localized signal may result from the cleaved N-terminal fragment or full length MYRF. To further clarify whether the full-length MYRF is trafficked out to the apical plasma membrane, we leverage a dual-tagged N-terminal FLAG and C-terminal MYC tagged construct (FLAG-MYRF-MYC). As expected, FLAG-MYRF which primarily tracks the N-terminal cleaved fragment, localizes to the nucleus of the RPE (Figure 4D and E), consistent with previous overexpression studies of MYRF. In contrast to our endogenous MYRF staining (Figure 4B and C), we observed little to no apical (Figure 4D, marked by phalloidin) or plasma membrane (Figure 4E, marked by NaK ATPase) localization of FLAG-MYRF. One potential explanation is that higher levels of MYRF through overexpression leads to increased trimerization and drives faster turnover (more cleavage) which limits our ability to capture the apically localized MYRF.

**Figure 4.**
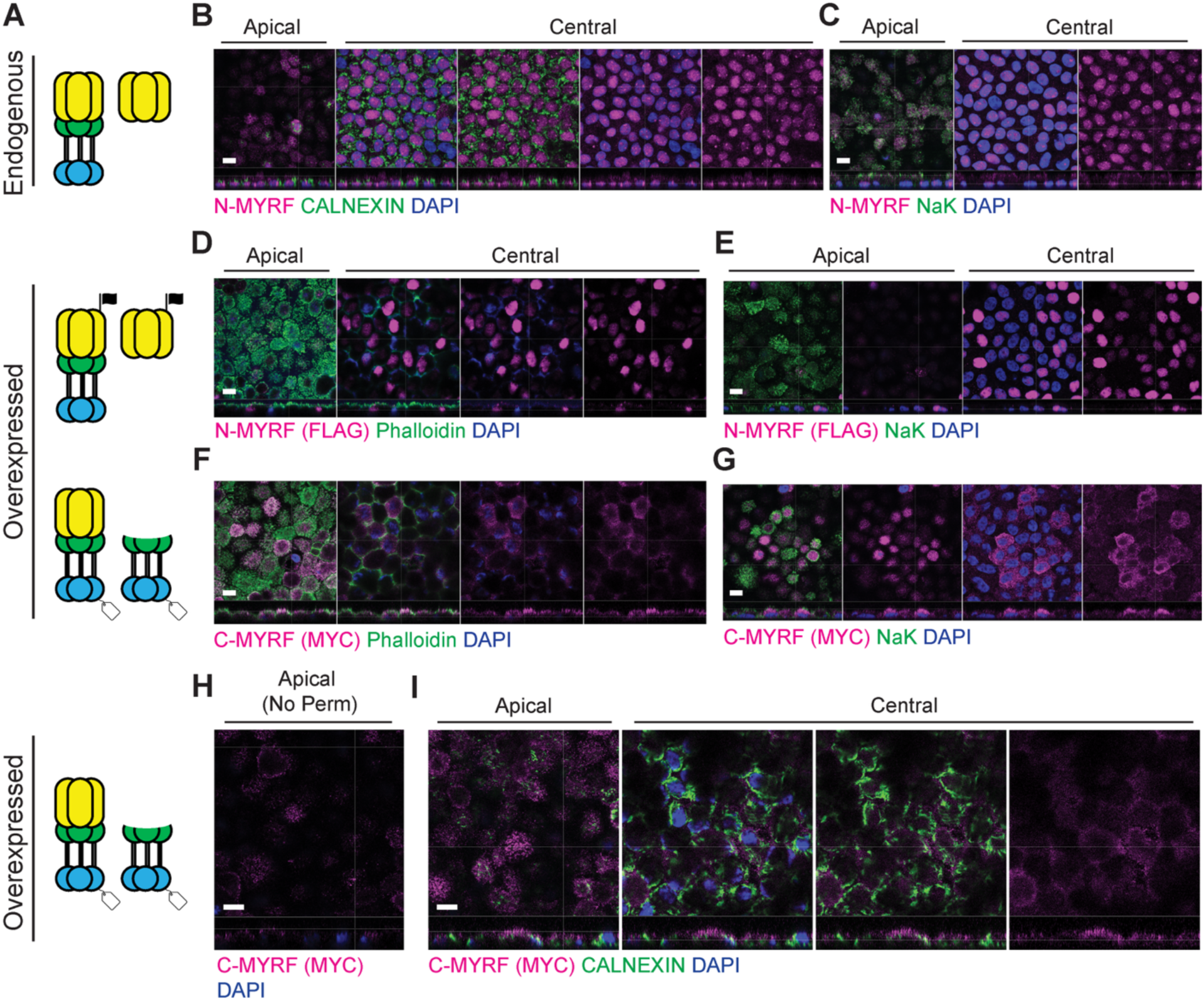
Overexpressed and endogenous MYRF are not exclusively ER or nuclear localized in hRPE. Schematic of MYRF fragments captured with each stain **(A).** Endogenous MYRF staining is both apical and nuclear localized showing overlap with nuclear marker DAPI **(B)** and plasma membrane marker NaK ATPase **(C)**. Endogenous MYRF does not show significant co-localization with ER marker Calnexin in mature hRPE **(C)**. Dual tagged FLAG-MYRF-MYC construct transduced into hRPE to determine localization of overexpressed N-MYRF and C-MYRF fragments. Immunofluorescent staining reveals FLAG-MYRF (N-MYRF) localizes to the nucleus as expected and shows little to no overlap with apical marker phalloidin **(D)** or the plasma membrane marker NaK ATPase **(E).** MYRF-MYC (C-MYRF) localizes to the apical and cytoplasmic compartments showing overlap with the apical marker phalloidin **(G)** and the plasma membrane marker NaK ATPase **(F).** MYRF-MYC (C-MYRF) localizes to the apical surface of non-permeabilized and unbleached hRPE (H) and does not primarily localize to the ER (Calnexin) in unbleached hRPE (I) (n=2-4 donors for each experiment). Scale bar = 10µm.

Meanwhile, we observed apical and cytoplasmic localization of MYRF-MYC (Figure 4F and G). MYRF-MYC signal co-localized with phalloidin (Figure 4F) and NaK ATPase (Figure 4G), suggesting that MYRF (either cleaved C-terminus or full-length) may be apically distributed to the plasma membrane in hRPE (Figure 4F). Furthermore, due to the orientation of MYRF, if it was localized to the plasma membrane we would expect to be able to detect the C-terminal portion on the cell surface, whereas the N-terminus would be intracellular. To strengthen our assertion that MYRF is localized to apical plasma membrane in hRPE, we showed that MYRF-MYC is detectable on the cell surface (Figure 4H) in non-permeabilized hRPE.

Lastly, we observed that full length cleavage deficient variants of MYRF (Figure 3), endogenous MYRF (Figure 4B and C), and FLAG-MYRF-MYC (Figure 4D-G) do not seem to be restricted to the ER in hRPE as previous studies have shown in oligodendrocytes. To evaluate the fraction of MYRF localized to the ER in hRPE, we co-stained cells transduced with FLAG-MYRF-MYC with ER marker CALNEXIN and MYRF-MYC. Our results show mostly diffuse cytoplasmic staining of MYRF-MYC with limited overlap between CALNEXIN and MYRF-MYC (Figure 4I). Together, our results support that MYRF (full length and/or the C-terminal cleaved product) are not solely retained in the ER in hRPE and instead localized to the apical plasma membrane.

### Inhibition of ER to Golgi Trafficking Increases Turnover and Cleavage of Endogenous MYRF in hRPE

To establish whether endogenous MYRF is localized to the ER and if localization to the plasma membrane is necessary for cleavage in the RPE, we treated hRPE with ER-to-Golgi trafficking inhibitor Brefeldin A (BFA) ^25,26^. Without BFA treatment, N-terminal MYRF, shows a diffuse nuclear distribution interspersed with small puncta. After 24 hours of BFA treatment, the mean signal intensity of nuclear MYRF was substantially decreased across three independent donors (Figure 5A, Supplemental Figure 1A, and Supplemental Table 1). Additionally, although trafficking of MYRF was blocked, we did not observe any accumulation of MYRF within the ER (Figure 5A). This may indicate that BFA is altering the cleavage dynamics and/or trafficking of endogenous MYRF in the RPE. We then performed Western blots on hRPE cell lysates treated with BFA for 24 hours, which revealed a decrease in full-length MYRF (0.59±0.45 FC vs. 1.0±0.27 FC in the control, p = 0.0185) and concomitant increase in the amount of cleaved N-MYRF (1.6±0.35 FC vs. 1.0±0.38 FC in the control, p = 0.0009) (Figure 5B and C). These data suggest that inhibition of trafficking by BFA treatment leads to faster turnover and cleavage of MYRF, but without ER retention.

**Figure 5.**
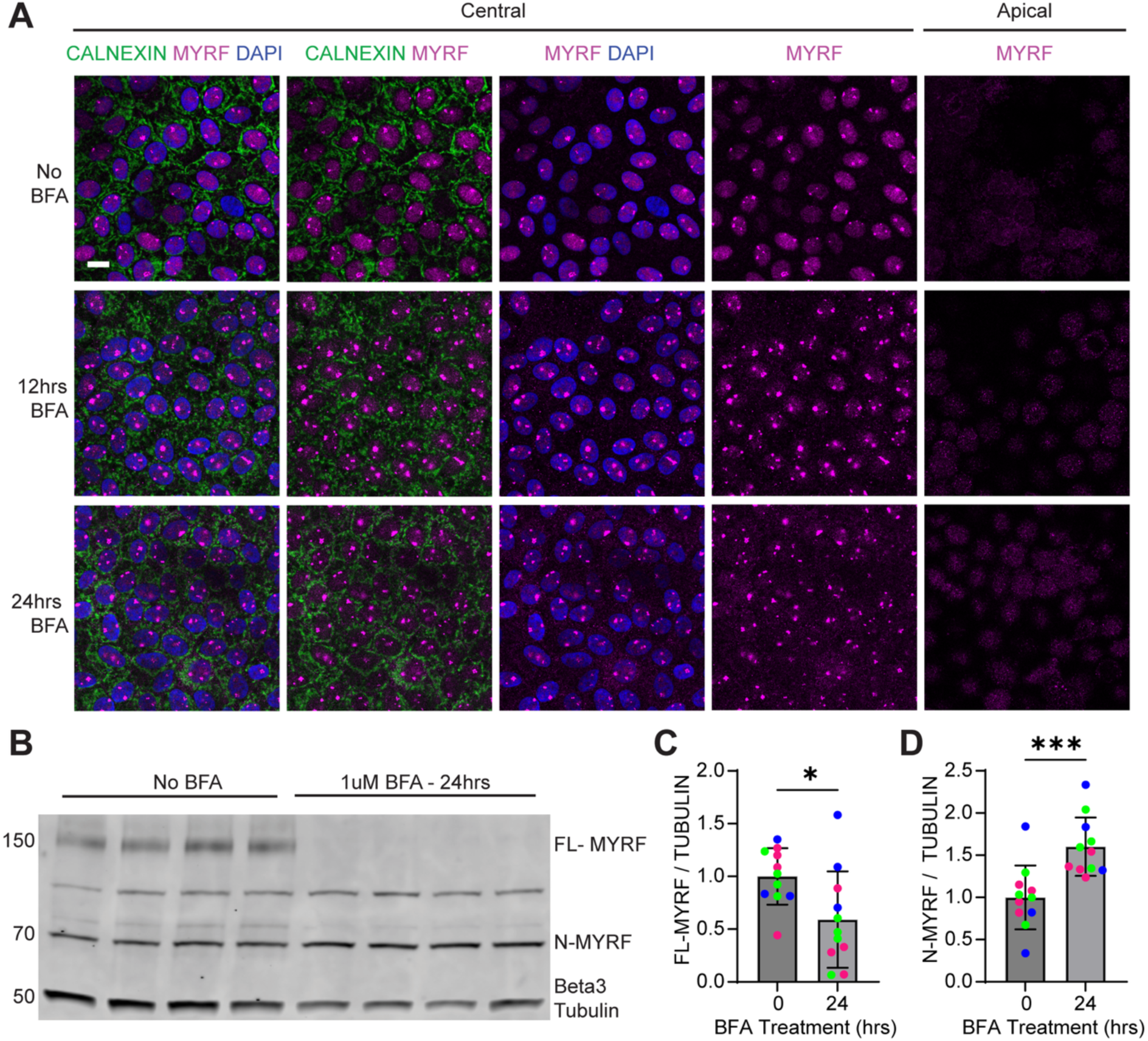
BFA treatment increases turnover and cleavage of MYRF in hRPE. **(A)** Time course of immunofluorescent staining of endogenous MYRF in BFA treated hRPE for 0, 12 or 24hrs. Endogenous apical and nuclear MYRF signal is decreased at the 12 and 24 hour time points (n=3 donors). **(B)** Western blot demonstrating endogenous MYRF has altered cleavage dynamics after inhibition of the ER to Golgi trafficking pathway (n=2 donors, each lane representing n=4 technical replicates per donor). **(C-D)** Quantification of full length and N-terminal MYRF protein normalized to beta-3 tubulin from western blot (n=3 donors, with n=4 technical replicates graphed per donor, each color represents a different donor). FL-MYRF decreased and N-MYRF increased following BFA treatment. **, p < 0.01; *, p <0.05, Student’s t-test. Scale bar = 10µm.

### Cleavage Deficient Variants use Classic ER to Golgi Trafficking to Reach the Apical Plasma Membrane

As WT MYRF appears to rely on canonical ER-to-Golgi pathways for proper cleavage dynamics (Figure 5), we next tested whether cleavage-deficient variant proteins are trafficked in a similar fashion despite mislocalization to the apical plasma membrane. We transduced hRPE with V679A-MYRF or R695C-MYRF. At baseline, the overexpressed variant proteins accumulate in the apical plasma membrane (Figure 3 and Figure 6A and B). Upon BFA treatment, however, cleavage-deficient variant proteins exhibited little-to-no change in cytoplasmic localization – and no colocalization with ER marker Calnexin (Figure 6A and B) - in a manner reminiscent of endogenous MYRF (Figure 5). Interestingly, after 24 hours of BFA treatment, the levels of apically localized V679A-MYRF and R695C-MYRF were reduced (Figure 6A and B). Western blots additionally revealed that total V679A-MYRF (0.16±0.05 FC vs. 1.0±0.11 FC in the control, p < 0.0001) and R695C-MYRF (0.11±0.08 FC vs. 1.0±0.17 FC in the control, p < 0.0001) protein levels were significantly reduced in BFA-treated hRPE as compared to untreated hRPE. Overall, our data suggest that cleavage-deficient variant proteins, like endogenous MYRF, use classic ER-to-Golgi trafficking mechanisms to reach the apical plasma membrane, and that protein levels are reduced in the presence of BFA.

**Figure 6.**
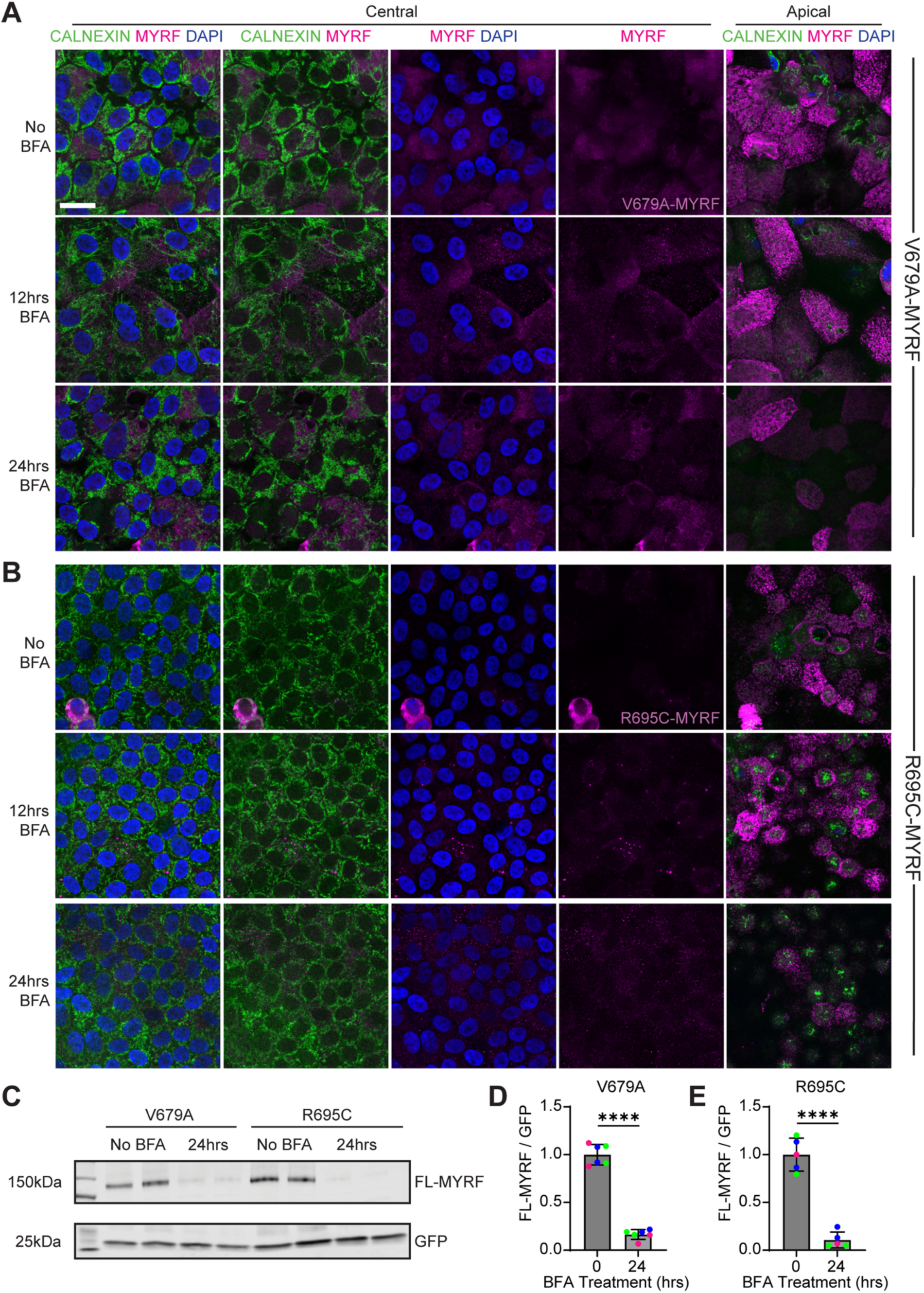
BFA treatment reduces protein levels of cleavage deficient MYRF variants. **(A)** Time course of immunofluorescent staining of V679A-MYRF in BFA (1uM) treated hRPE for 0, 12, and 24hrs. Protein level of V679A-MYRF appears to be reduced apically after 24hrs (n=3 donors). **(B)** Time course of immunofluorescent staining of R695C-MYRF in BFA (1uM) treated hRPE for 0, 12, and 24hrs. Protein level of R695C-MYRF appears to be reduced apically after 24hrs (n=2 donors). Scale bar = 20µm. **(C)** Western blot of hRPE lysates transduced with V679A or R695C-MYRF untreated or with 1uM BFA treatment for 24hrs. Both V679A **(D)** and R695C-MYRF **(E)** protein levels are reduced as quantified relative to GFP (n=3 donors, with n=2 technical replicates graphed from each donor and each color represent a different donor).*, p <0.05 by Student’s t-test.

## DISCUSSION

We provide multiple lines of population (PM2), computational (PP2, PP3, PM5) and functional (PS3) evidence demonstrating that *MYRF* p.R695C – identified in an adult patient with nanophthalmos – is pathogenic according to the American College of Medical Genetics (ACMG) criteria (Table 1) ^27^. Furthermore, in characterizing the molecular consequences of this variant, we reveal critical insights into the regulation and trafficking of MYRF in polarized hRPE. Our study thus demonstrates the utility of studying rare, disease-associated variants in advancing our knowledge of basic protein biology and function.

### Phenotypic Spectrum of Variants in MYRF

Variants in *MYRF* contribute to a spectrum of human disease ranging from severe syndromic presentations to isolated ocular manifestations ^13^. Curiously, even variants within the same functional domain can have differing disease mechanisms (haploinsufficiency vs. dominant negative) with variable impact across different organ systems ^10,14^.

In this study, the patient’s features fit with classic nanophthalmos with the exception of macular yellow deposits (first observed in her 30s), which may represent early stages of subretinal disruption or a recently described macular phenotype termed Benign Yellow Dot Maculopathy ^28^. These macular deposits and OCT evidence of mild subretinal discrete irregularity in the outer retina and RPE are not a common finding in nanophthalmos patients and point to potential RPE dysfunction. To our knowledge, the p.R695C variant represents the first reported association between *MYRF* and significant macular fundus and OCT findings, and this disease mechanism appears to be different from our previously described C-terminal variant in *MYRF*, p.G1126fs*30, which was dominated by RPE developmental defects, no signs of MYRF mislocalization, and no evidence of macular deposits or discrete outer retinal/RPE disruption in the macula ^5–7^.

Interestingly, the isolated ocular presentation associated with *MYRF* p.R695C is a notable contrast from the syndromic presentation reported for a different substitution at the same amino acid residue, p.R695H ^11^. One explanation for the phenotypic variability between p.R695C and p.R695H could be a differential tolerance of the MYRF trimer to different variant proteins. Since MYRF functions as a homotrimer, a heterozygous variant does not necessarily lead to a 50% loss of function. From an equal pool of wild-type (WT) and variant subunits, random assembly yields eight possible combinations across four trimer types: 12.5% all-WT, 37.5% with two WT subunits (and one variant), 37.5% with one WT subunit (and two variants), and 12.5% all-variant ^11^. Consequently, if a trimer can tolerate up to two variant copies, functional trimers (containing 3, 2, or 1 WT subunits) would total 87.5%, resulting in only a 12.5% loss of function and leading to only ocular disease. In contrast, a variant that tolerates only one variant copy (ex. p.R695H) retains 50% functional trimers, resulting in the expected 50% loss of function ^11^. Further experiments would be needed to support this speculation.

### MYRF p.R695C Variant is Cleavage Deficient and Avoids ER Retention

MYRF is a transcription factor that requires a multistep process to initiate gene activation. First, MYRF must homotrimerize at the ER membrane. It must then undergo cleavage to release an N-terminal fragment, which translocates to the nucleus and activates gene expression at target sites. *In silico* modeling of the p.R695C variant using a publicly available MYRF crystal structure suggested the same losses in hydrogen bonding thought to ablate cleavage of MYRF p.R695H, indicating similar molecular mechanisms across both variants. By demonstrating cleavage deficiency in both ARPE-19 and polarized primary RPE cultures we established that impaired autoproteolysis is an intrinsic property of the variant rather than a cell-line artifact. Furthermore, the absence of ER accumulation challenges the prevailing model derived from oligodendrocytes and suggests that MYRF trafficking is fundamentally influenced by epithelial polarity. While these results were unexpected, they are not entirely surprising considering that no ER retention sequence has been identified or predicted in MYRF. It may be that full-length MYRF does not remain stagnant at the ER membrane, and that processing/trafficking of MYRF is cell-type specific. One intriguing possibility is that MYRF is being retained or trafficked in other membrane-bound vesicles within the RPE (Figure 7), but additional studies are needed to support this hypothesis.

**Figure 7.**
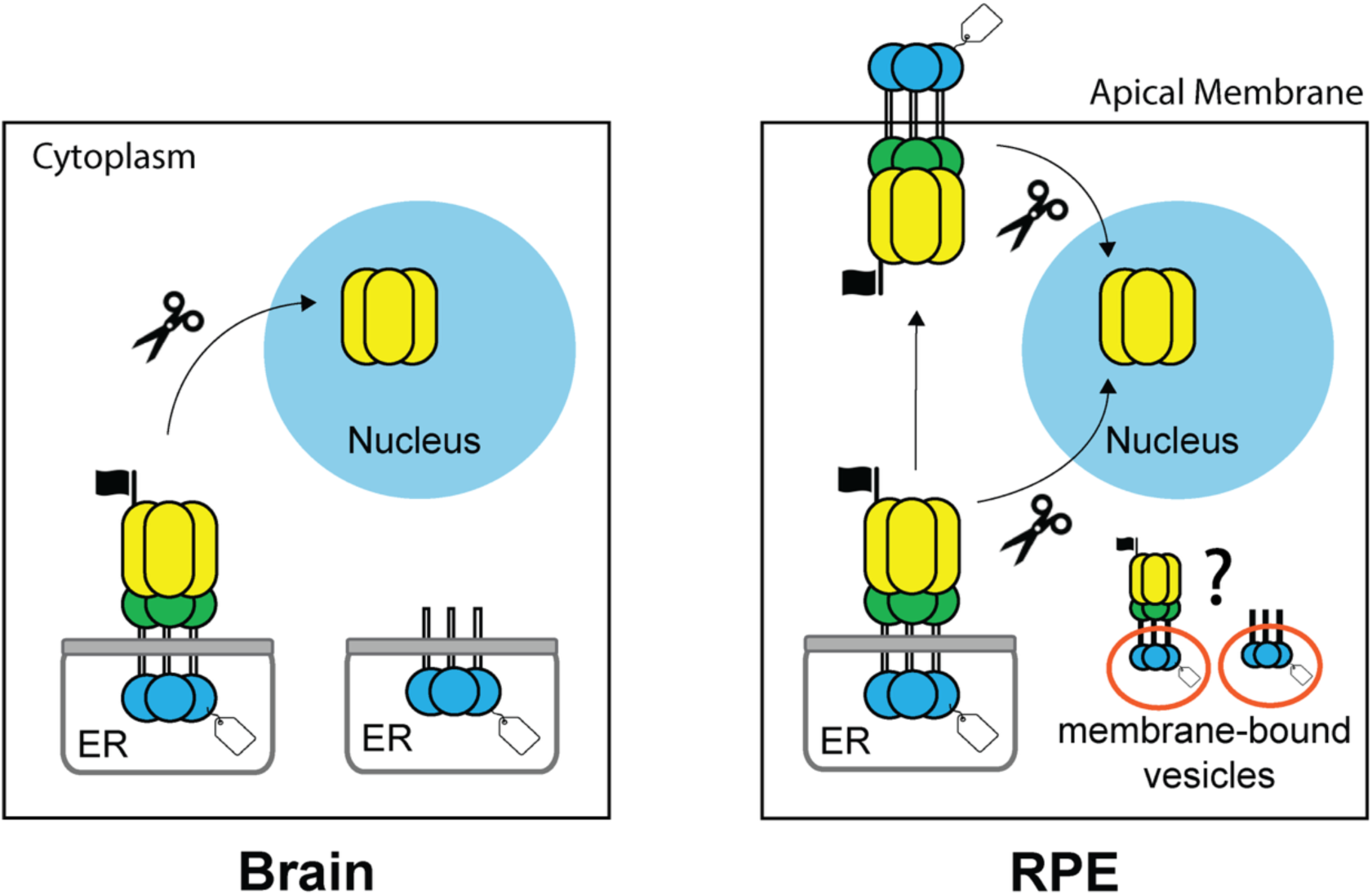
Model for MYRF trafficking in the RPE. Normal MYRF trafficking in the RPE is not restricted to the ER and likely resembles the mechanisms previously described in *Dictyostelium discoideum* and *C. elegans* whereby MYRF is trafficked to the plasma membrane as a cleavage regulatory mechanism. The lack of ER retainment, presence of a transmembrane domain in MYRF, and diffuse cytoplasmic localization of the full length and C-terminal fragment of MYRF open the possibility the MYRF may be present in membrane-bound vesicles within hRPE.

### Cell-Type Specific Function and Processing of MYRF

*MYRF* is expressed across many different organs, tissue types, and it is highly conserved across species ^13^. Its function and processing have been most notably described in the development/maturation of oligodendrocytes ^8,24,29^, RPE ^5–7^, lung mesothelial cells ^30^, and the heart ^31^ in mammalian cells *in vitro* or *in vivo* in mice. Homologs of MYRF have also been functionally characterized such as MYRF-1 in the synaptic rewiring of dorsal neurons in *C. elegans* ^22^ and MrfA regulating *Dictyostelium* prestalk cell differentiation ^32^. Altogether MYRF has an evolutionarily conserved role in development, differentiation, and transcriptional programming across organs and cell types.

Key processing steps of MYRF remain largely conserved across species and cell types including: trimerization, cleavage, and nuclear translocation to activate gene expression. For example, the MrfA homolog undergoes trimerization, cleavage, and nuclear translocation in *Dictyostelium* and this is conserved in mouse oligodendrocytes^8^ and in primary human RPE cell cultures ^7^. However, studies in *C. elegans*, *Dictyostelium,* and mice suggest that the mechanisms that regulate these processing steps may vary across species and developmental timing. Our study provides further evidence to support this idea and demonstrates that the RPE may have their own unique mechanisms for regulating MYRF processing.

Here we present compelling evidence that MYRF (full length and/or its cleaved fragments) localizes to the plasma membrane in human RPE (Figure 3 and Figure 4), which has not been observed in any other mammalian cell type. Interestingly, in *C. elegans* MYRF-1 is also trafficked to the plasma membrane, however it’s localization to the plasma membrane and subsequent cleavage is regulated by its partner protein, P-granule-associated novel protein 1, PAN-1. While PAN-1 does not have a human homolog, one potential PAN-1 like regulator of MYRF plasma membrane localization in human RPE is Transmembrane protein 98, TMEM98. Variants in TMEM98 are associated with nanophthalmos ^4,33^ and it acts as an inhibitor of MYRF cleavage ^6,34^. Furthermore, TMEM98 is localized similarly to MYRF in mouse RPE showing ER, diffuse cytoplasmic, and apical localization ^6,34^. It is tempting to speculate that TMEM98 could be influencing apical localization and processing of MYRF in RPE, but more work remains to determine what role, if any, TMEM98 plays in regulating MYRF localization.

Our studies using BFA provide some insights into the trafficking mechanisms that govern MYRF movement within the RPE. Our characterization of endogenous MYRF in the presence of BFA demonstrated decreased nuclear localization, an increased concentration of N-terminal cleavage fragments, and no accumulation within the ER. These results suggest that MYRF relies on ER-to-Golgi trafficking for proper nuclear localization and cleavage dynamics. Additionally, our observation of reduced total levels of cleavage-deficient MYRF following BFA treatment indicates that these variant proteins rely on similar mechanisms to reach the plasma membrane. Treatment with BFA also reduced overall levels of full-length MYRF in all conditions. This raises the possibility that BFA may be inducing ER stress, activating the unfolded protein response (UPR), and inhibiting protein synthesis within the hRPE cultures. Additional studies are needed to experimentally validate these results. Still, these data support that MYRF is not retained by the ER in the RPE as inhibition of trafficking did not result in ER accumulation. Altogether, it appears that both wildtype and cleavage-deficient MYRF rely on classic ER-to-Golgi trafficking for proper processing and function (Figure 7B), and that the lack of ER accumulation is likely due to fast turnover and/or inhibition of protein synthesis induced by the UPR. However, Golgi-independent trafficking of MYRF in the RPE cannot be definitively ruled out with our available data.

### Conclusions

In this study, we investigated the molecular mechanisms of the *MYRF* ICA domain variant p.R695C – identified in a patient with nanophthalmos and RPE/subretinal irregularities – using *in silico* modeling and *in vitro* functional assays with human RPE. Our results emphasize the importance of studying protein processing and function in not only a cell-type-specific manner, but in well-polarized and differentiated cells that can mimic *in vivo* conditions. Using human RPE-like cell lines, we did not observe the apical mislocalization that was obvious in mature, polarized hRPE. We also show that wildtype and cleavage-deficient MYRF may rely on ER-to-Golgi trafficking for proper localization and cleavage dynamics. These results build upon the mounting evidence that MYRF processing and function are regulated in a tissue-specific manner, which may reflect the differences in MYRF-mediated transcriptional regulation across various cell types. Moreover, we demonstrate that, in RPE, MYRF is localized to the plasma membrane, and processing mechanisms may be evolutionarily conserved from lower order organisms such as *C. elegans*. Given the broad expression and key roles of *MYRF* in multiple organs, future work is needed to understand how function and processing of MYRF vary across different cell types.

## METHODS

### Patient Collection, Sequencing, and Analysis

Patient saliva samples were collected, extracted, and sequenced as previously described^7,35^. Sanger sequencing was performed (Eurofin Genomics) for MYRF exon 15 and 16 to evaluate segregation in unaffected children using forward: 5’-tgacatctgagacaccagtc-3’ and reverse: 5’-aagaaagagaaacggggctg-3’ primers ^6^.

### *In Silico* Structural Modeling

*In silico* modeling of the amino acid substitution to evaluate changes in amino acid interactions was done by mutating the crystal structure PBD file (pdb_00007dc3) ^12^ in PyMOL.

### ARPE-19 and HEK293T Studies

#### Protein Localization Studies

ARPE-19 cells were seeded at 40,000 cells/well on Nunc Lab-Tek II 4-well chamber slides. After 24 hours, cells were transfected with 1500ng of WT-MYRF, V679A, or R695C plasmid DNA using the ViaFect™ Transfection Reagent (Promega, E4981, Madison, WI) at a 6:1 (ViaFect:DNA) ratio per well. Transfected ARPE-19 cells were stained and imaged as previously described ^7^.

#### Protein Cleavage Studies

ARPE-19 cells (ATCC, #CRL-2302) were seeded at 40,000 cells in 24-well tissue culture plates. After 24 hours, sub-confluent cells were transduced with lentiviral vectors carrying WT-MYRF, V679A, or R695C (MOI 3 from 10X concentrated stocks) vectors in the presence of 1mg/mL LentiBoost (Mayflower Bio, # SB-P-LV-101-11). Whole cell lysates were collected for western blotting as previously described ^7^. PVDF membranes were probed chicken anti-GFP 1:5000 (Abcam, #13970), rabbit anti-FLAG 1:1000 (CST, #14793), and/or rabbit anti-MYRF (gift from Ben Emery) primary antibodies. IRDye 800CW goat anti-rabbit IgG 1:20,000 (LICOR, 926-32211) and IRDye 680RD donkey anti-chicken IgG 1:20,000 (LICOR, #925-68075) were used as secondary antibodies. Blots were imaged using the Odyssey CLx (LICOR, Lincoln, NE) and quantified using ImageStudio.

#### HEK293T Synthetic Luciferase Reporter Assays

HEK293T cells (ATCC, #CRL-3216) were seeded at 200,000 cells in 24-well tissue culture plates. After 24 hours, sub-confluent cells were transfected using the Lipofectamine 2000 (Thermo Fisher, #11668027) with 500ng of total DNA vectors. Conditions and DNA amounts listed in Supplemental Table 2. Cells were incubated with transfection reagents for 6 hours and then removed. After 96 hours, cell lysates were collected and luciferase activity was measured according to the Dual-Luciferase® Reporter Assay System. Cells were collected in 100uL of 1X Passive Lysis Buffer and passively lysed on a shaking platform for 15 minutes, and after 20uL were transferred into a white 96-well flat bottom plate (#3912, Corning). The plate was then run on a Promega GloMax 96 Instrument (Model #E6521, Promega). Luciferase activity levels were normalized to transfection efficiency via TK Renilla (Firefly Luciferase Activity / TK Renilla activity) and fold change in activity levels were normalized to cells transfected with Empty Vector only.

### hRPE Studies

#### Transduction of Lentiviral Vectors in hRPE

Mature hRPE grown for two months on transwell inserts were transduced as previously described ^7^ with WT-MYRF, V679A, or R695C (MOI 5 from 10X concentrated stocks) vectors in the presence of 1mg/mL LentiBoost (Mayflower Bio, # SB-P-LV-101-11). All primary cells were genotyped for Age Related Macular Degeneration risk SNPs as previously described ^7^.

#### Protein Localization Studies

Protein localization studies in hRPE were performed as previous described ^7^. After non-specific blocking, cells were incubated with various combinations of anti-FLAG 1:1000 (CST, #14793), anti-MYC (1:400), anti-Calnexin 1:200 (Sigma, #C4731), anti-Na^+^/K^+^-ATPase 1:1000 (Santa Cruz Biotechnology, #sc58628) and/or anti-GFP (1:1000), primary antibodies for 1 hour at RT. Fixed cells were then incubated with anti-goat rabbit Alexa Fluor 647 1:1000 (Invitrogen, #A-21446), goat anti-mouse Alexa Fluor 594 1:1000 (Invitrogen, #A-11005), goat anti-mouse Alexa Fluor 555 1:1000 (Invitrogen #A32727), donkey anti-chick Alexa Fluor 488 1:1000 (Jackson ImmunoResearch, #703-545-155), and/or phalloidin Alexa Fluor 568 (Invitrogen, #A12380) secondary antibodies for 1 hours at RT. Transwell membranes were mounted and imaged as previously described^7^. For cell surface staining, cells followed all the same steps except they did not undergo a permeabilization step, and all other steps were performed in a blocking buffer without permeabilizing agent.

#### Protein Cleavage Studies

Mature hRPE grown on 96-well plates were transduced as described in the “Transduction of Lentiviral Vectors in hRPE” section and cell lysates collected for western blotting as previously described ^7^. Protein quantification, denaturation, SDS-page, and imaging was performed as described for ARPE-19 cells above.

#### Transepithelial Electrical Resistance (TEER) Measurements

TEER was measured as previously described ^19,20^ with a volt-ohmmeter and electrode probe (EVOM and STX2; World Precision Instruments, Sarasota, FL, USA).

#### Inhibition of ER Protein Transport via Brefeldin A in hRPE

hRPE were transduced as described in the “Transduction of Lentiviral Vectors in hRPE” section. One week post-transduction, transduced and non-transduced hRPE were treated with 1uM of Brefeldin A for 0, 12 or 24 hours to inhibit ER to Golgi protein transport. After treatment with Brefeldin A, cells were fixed, bleached, and blocked according to above methods (see <u>Protein Localization Studies</u>). After blocking, hRPE were incubated with either anti-FLAG 1:1000 (CST, #14793), anti-MYRF 1:500 ^23^, anti-and/or Calnexin 1:200 (Sigma, #C4731) primary antibodies for 1 hour at RT. Fixed cells were then incubated with anti-goat rabbit Alexa Fluor 647 1:1000 (Invitrogen, #A-21446) and goat anti-mouse Alexa Fluor 594 1:1000 (Invitrogen, #A11005) secondary antibodies for 1 hours at RT. Transwell membranes were mounted and imaged as described in the “Protein Localization Studies” section.

### Statistics

Prism 10 was used to visualize and perform statistical analysis on quantitative datasets. One-way ANOVA was used when comparing the means of 3 or more groups. Student’s t test was used when comparing the means of only 2 groups. Unless otherwise noted, in the figure legend data represent mean ± SD.

### Study Approval

Human subjects work was approved by the Institutional Review Board (IRB) at the University of Michigan under protocol HUM00046159. All subjects provided written informed consent for their inclusion. This study was carried out in accordance with the Tenets of the Declaration of Helsinki. Patient and sample identifiers are only known to the study team and cannot otherwise be relinked.

## Data Availability

All additional relevant data can be found within the article, and its supplementary information and primary data publicly available. Genomic data will be accessible at dbGAP (accession no. phs004462.v1.p1).

## Author Contributions

GMR and LP wrote the manuscript and designed research studies. GMR and RL conducted experiments, acquired data, analyzed data. BB collected patient data and DNA samples. LP, JYSH, JMLM, RBH, RY, and SEM, TEH provided reagents. BG, WP, EU, and AN analyzed data. All authors reviewed and revised the manuscript.

## Acknowledgements

We thank our funding including NEI K08 EY032098, the E. Matilda Ziegler Foundation for the Blind, Bright Focus Foundation (M2022011N), Research to Prevent Blindness Career Development Award, and the Knight Templar Eye Foundation supporting to L.P., NEI P30-EY007003, Shared Instrument Grant S10OD28612, NEI T32 EY013934 (G.R.), James Grosfeld Initiative for Dry AMD Trainee Grant (G.R.), NEI intramural funding (R.B.H., B.G.), NEI K08 EY033420, Discovering Hope Foundation, and James Grosfeld Initiative for Dry AMD support to J.M.L.M.; BrightFocus Foundation M2024001F and the Michigan Pioneer Fellows Program to J.Y.S.H. We also thank members of the Miller lab (Gillian Gulette and Qitao Zhang) for their contribution of human primary cells, protocols, and expertise, Enrique Rodriguez-Boulan lab for the RPE bleaching protocol, Ben Emery for providing reagents and guidance on the project, Michael Wegner for generously providing the luciferase assay constructs, Giuseppe R. Diaferia and Gioacchino Natoli for the endogenous N-terminal MYRF antibody, and Sally Camper for helpful insights and critical reading of the manuscript. No federal funds were used for HFT research. This research was supported in part by the Intramural Research Program of the National Institutes of Health (NIH), The National Eye Institute. The contributions of the NIH authors were made as part of their official duties as NIH federal employees, are in compliance with agency policy requirements, and are considered Works of the United States Government. However, the findings and conclusions presented in this paper are those of the authors and do not necessarily reflect the views of the NIH or the U.S. Department of Health and Human Services. This work is the result of NIH funding, in whole or in part, and is subject to the NIH Public Access Policy. Through acceptance of this federal funding, the NIH has been given a right to make the work publicly available in PubMed Central.

## SUPPLEMENTAL TABLES

**Supplemental Table 1.**
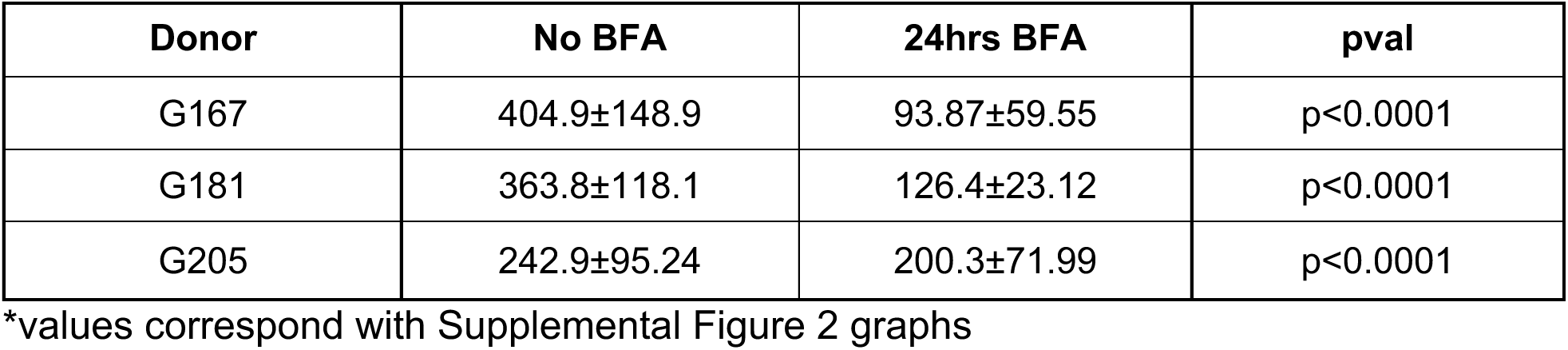
BFA Treatment Significantly Reduces Mean Nuclear Intensity of Endogenous MYRF.

**Supplemental Table 2.** Synthetic Luciferase Reporter Assays Transfection Reactions.

| **Plasmids** | **Empty** | **WT-MYRF** | **R695C-MYRF** | **SOX10** | **SOX10 + WT-MYRF** | **SOX10 + R695C-MYRF** |
|---|---|---|---|---|---|---|
| SOX10 | - | - | - | 66 | 66 | 66 |
| WT-MYRF | - | 264 | - | - | 264 | - |
| R695C-MYRF | - | - | 264 | - | - | 264 |
| Empty | 330 | 66 | 66 | 264 | - | - |
| dimluc c7c<br>(SOX10 + MYRF synthetic reporter)<sup>18</sup> | 165 | 165 | 165 | 165 | 165 | 165 |
| TK renilla | 5 | 5 | 5 | 5 | 5 | 5 |
| Total DNA per reaction (ng) | 500 | 500 | 500 | 500 | 500 | 500 |

## SUPPLEMENTAL FIGURES

**Supplemental Figure 1.**
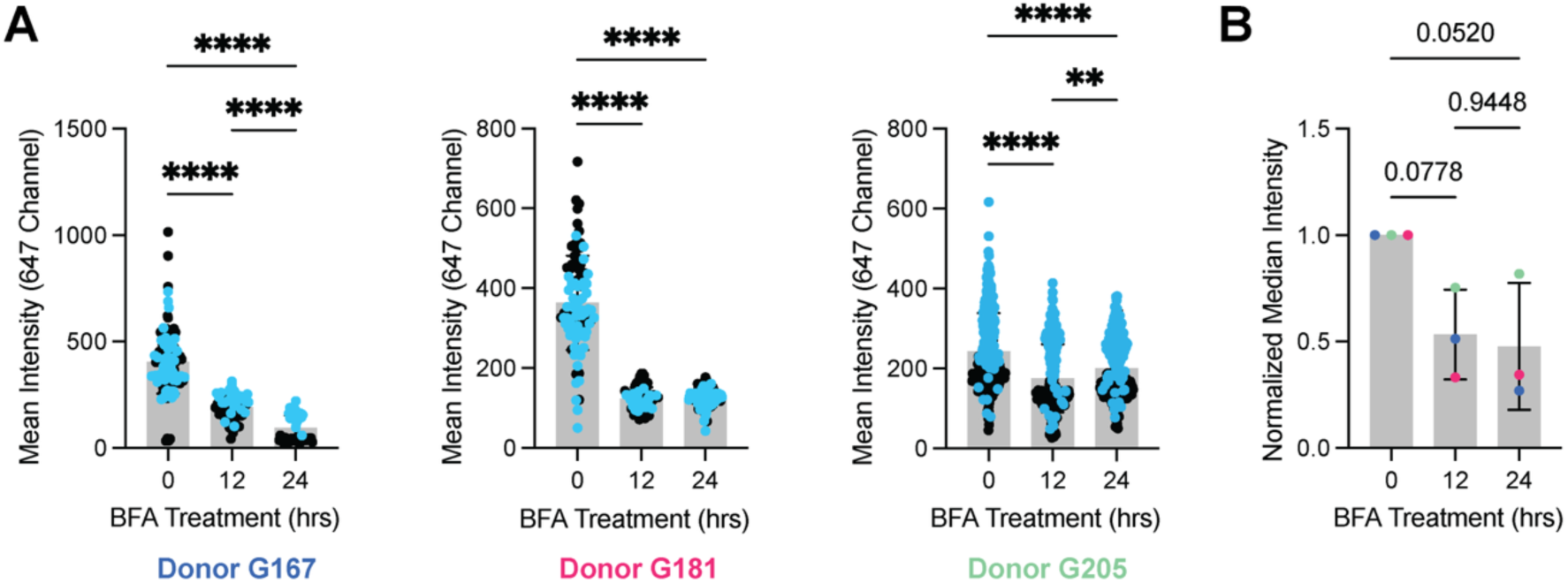
Quantification of endogenous MYRF nuclear signal in the presence of BFA. **(A)** Mean intensity of endogenous MYRF (647 channel) within the nuclear surface decreases overtime with BFA treatment. Blue dots represent technical replicate 1 and black dots represent technical replicate 2, (n=3 donors). **(B)** Normalized median intensity of endogenous MYRF showing reduced median nuclear signal intensity of endogenous MYRF across donors (blue = G167, pink = G181, green = G205). One way ANOVA was used to determine statistical significance. ****, p < 0.0001; **, p < 0.01; *, p <0.05.

